# Remote patient monitoring and digital therapeutics in heart failure: lessons from the Continuum pilot study

**DOI:** 10.1101/2022.12.06.22283117

**Authors:** Emmanuel Marier-Tétrault, Emmanuel Bebawi, Stéphanie Béchard, Philippe Brouillard, Priccila Zuchinali, Emilie Remillard, Zoé Carrier, Loyda Jean-Charles, John Nguyen, Pascale Lehoux, Marie-Pascale Pomey, Paula A. B. Ribeiro, François Tournoux

**Affiliations:** University of Montreal, Montreal, QC, Canada; Hospital of the University of Montreal (Centre Hospitalier de l’Université de Montréal, CHUM) QC, Canada; @Coeur Research Lab - Research Center of the CHUM, Montreal, Quebec, Canada; Sacred Heart Hospital, Montreal, QC, Canada

**Keywords:** chronic heart failure, telemonitoring, digital therapeutics, software as medical device (SAMD), remote patient management, digital health, mobile health, algorithms

## Abstract

**Introduction:** The increasing use of digital health solutions to monitor heart failure (HF) outpatients has been driven by the COVID-19 pandemic. An ideal technology should answer the specific needs of a public healthcare system: easy integration and proof of clinical benefit to justify investment in its long-term use. Through a consortium bringing together patients, physicians, industry, and hospital organizations, we developed a digital solution called “Continuum,” targeting patients with HF and other comorbidities.

**Hypothesis:** A digital health solution combining remote patient monitoring (RPM) and digital therapeutics (DTx) was developed to ensure a better follow-up of patients and to rapidly optimize their medication and subsequently avoid future severe adverse events.

**Methods:** A pilot intervention/control study with a three-month follow-up was conducted. Patients in the intervention group (remote patient monitoring group, RPM^+^) had a smartphone or tablet and entered in their mobile app their vital signs, weight, and HF symptoms daily. HF patients who either did not have a mobile device or the skills to use the app were enrolled in the control group (RPM^-^). The HealthCare Professionals (HCPs) used a web-based dashboard to follow the RPM^+^ patients. They could access the results of a DTx solution to help them optimize the HF treatment according to Canadian guidelines.

**Results:** 52 HF patients were enrolled in this study, 32 in the RPM^+^: 69±9y age, 75% male, ejection fraction 42 ± 14%. In the RPM^-^ group, more patients had at least one hospitalization (all-cause) compared to the RPM^+^ group (35% versus 6% respectively; p=0.008). Similarly, the number of patients with at least one HF hospitalization was more significant in the RPM^+^ group compared to the RPM^-^ (25% versus 6%, p=0.054). Finally, the intervention showed a medium effect on HF treatment optimization (w=0.26) and quality of life for the most compliant patients to the intervention (g=0.48).

**Conclusion:** The results of this pilot study demonstrated the feasibility of an intervention combining RPM and DTx solutions for HF patients. Preliminary results suggest promising impacts on quality of life, hospitalizations, and patients’ medication optimization. However, they need to be confirmed in a more extensive study.

## Introduction

Heart failure (HF) is the leading cause of hospitalization in Canada’s patients over 65. (1) HF patients need frequent clinical follow-ups to prevent readmission and to have their medical therapy optimized according to national (2) and international guidelines. (3,4) Unfortunately, many barriers to implementing these guidelines exist, including limited human and financial resources, which explains part of the current therapeutic inertia and the recurrent adverse events in this population. (5,6)

The COVID-19 global pandemic has exacerbated this phenomenon (7) and forced the rapid development of digital health solutions to address this issue. (2,8) Many remote patients monitoring (RPM) tools developed for HF (9,10) are now implemented in the clinical routine. However, despite the growing adoption of these technologies by healthcare professionals, evidence is still scarce: the European Society of Cardiology made RPM a class 2B recommendation (11) while the American Heart Association did not even recommend it in their latest guidelines. (4) Besides RPM systems, new tools to encourage and support patient medical therapy optimization (best known as Digital Therapeutics (DTx) solutions) have been developed (12–14) and preliminary data suggest some benefits for HF patients. (15) In the specific context of HF, the main obstacle to the development of these DTx solutions is the presence of other comorbidities (such as diabetes, renal failure, or chronic obstructive pulmonary disease) which all have their guidelines on patient management. It adds extra layers of complexity when programming the DTx algorithms since these recommendations are not always perfectly aligned with each other.

The Continuum project is a joint initiative of a healthcare center, a software start-up, and an industrial partner to develop an intervention which combines an efficient RPM system and a DTx solution for HF. We present the pilot phase results, which aim to test the feasibility of implementing an innovative digital solution in a natural healthcare environment and assess its potential benefits to our HF patients with various comorbidities.

## Methods

### Study design

This prospective pilot study was conducted in our HF clinic at the Research Center of the Hospital of Montreal University (CRCHUM) and at the University of Montreal Hospital Center (CHUM) between October 2020 and June 2021. NYHA class, 2 to 4 HF patients (age≥18y) with diabetes were enrolled regardless of the value of their left ventricular ejection fraction (LVEF). Exclusion criteria were terminal kidney failure, cancer with a prognosis <1-year, severe cognitive impairment, terminal cirrhosis and patient at high risk of loss of follow-up (e.g. history of non-adherence to treatment or substance abuse limiting follow-up appointments).

Both RPM and DTx systems were tested during this pilot study. Patients were first allocated to the RPM^+^ group if they 1) owned a smartphone or a tablet and 2) could use our dedicated mobile application. The ability of the patient (or its caregiver) to use the app was tested by a nurse trained explicitly for this purpose. After an initial demonstration, the patient (or its caregiver) had 15 minutes to demonstrate their ability to use the app. All the patients who did not have a mobile device or failed the ability test were allocated to the RPM control group (RPM^-^). Patients were considered highly compliant with the intervention if they used the app >80% of the days during the 12 weeks of the study.

All the patients in the RPM^+^ group had their data analyzed by the DTx system (RPM^+^/DTx^+^ group) (figure 1). Patients in the RPM^-^ group were randomly divided into two subgroups (1:1): a subgroup called RPM^-^/DTx^+^ where the data were still analyzed by the DTx module even in the absence of the RPM app, and a subgroup for which no analysis by the DTx module was performed (RPM^-^/DTx^-^). This subdivision for the RPM^−^ patients was performed to assess the potential interest in using the DTx system alone. To summarize, three groups of patients were followed during this pilot study: RPM^+^/DTx^+^, RPM^-^/DTx^+^ and RPM^-^/DTx^-^.

**Figure 1:**
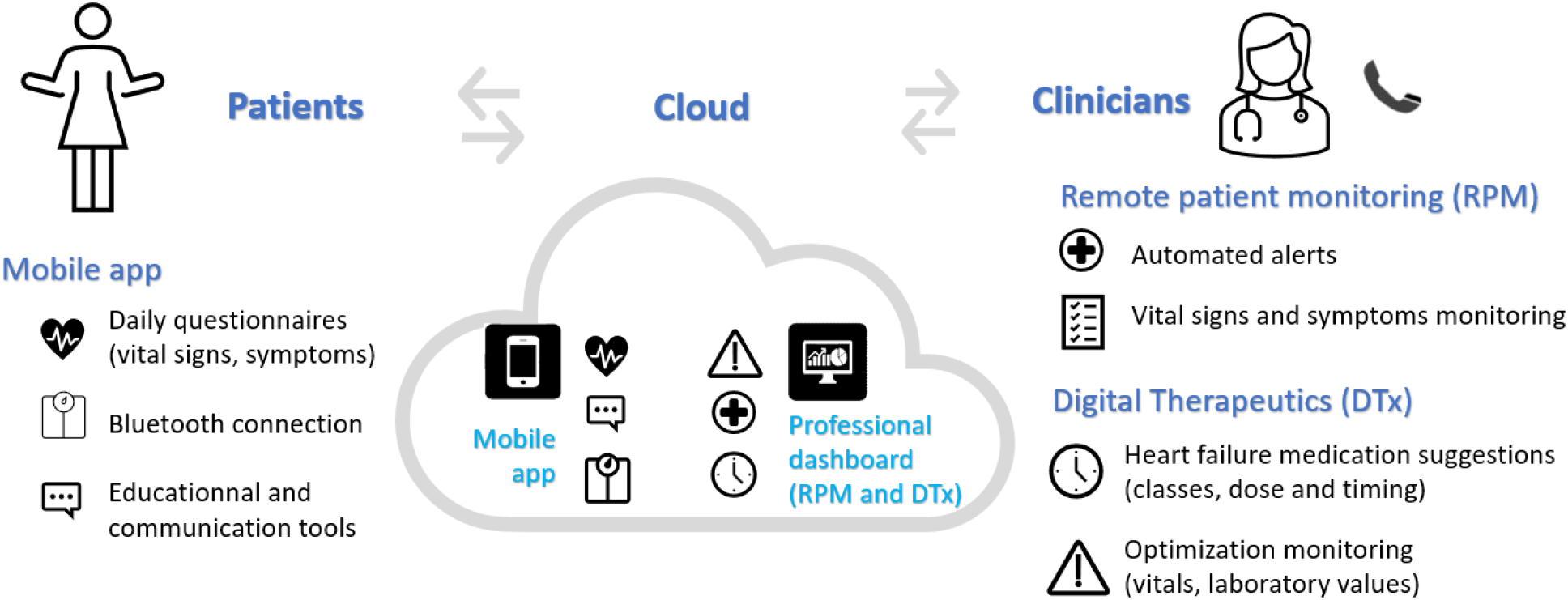
Mobile and professional interfaces of the application.

Each patient was then followed and assessed at three time points: enrolment (day 0), 6 and 12 weeks. Quality of life (Kansas City Cardiomyopathy Questionnaire-12 (KCCQ-12)), hospitalizations and HF medication were assessed for each visit. A patient with LVEF <40% was considered to have its HF medication optimized if a minimum of one drug class among those recommended by the Canadian guidelines was added to the patient’s profile or if the dose of one of its HF drugs was increased during the follow-up.

Our local ethics committee approved the study protocol (21.403), and all the patients gave their written consent to the study.

### The Continuum solution

Each RPM^+^ patient was offered a package with three Bluetooth-connected devices: a scale, a blood pressure monitor, and a glucometer. The use of these devices was not mandatory. The patient could decide to use one, two or all of them. Technical support was provided to help them connect these devices to their phone or tablet. Patients’ data were either automatically (through Bluetooth) or manually entered within the mobile app and transferred to the Continuum platform. These data included blood pressure, heart rate, weight, glycemia, number of steps, and answers to a short survey about HF symptoms. Specific educational contents (i.e., HF symptoms, nutrition, physical activity, and mental health) were accessible within the mobile app. The platform sent frequent reminders through the app to keep the patients engaged. Once a day, from Monday to Friday, a clinical nurse accessed the web-based dashboard (Figure 1) and reviewed the data and alerts triggered according to specific algorithms within our Continuum platform. Depending on the type of alert, the nurse could call the patient and perform a virtual visit or ask the patient to come for an urgent cardiology consult.

The DTx module was integrated into the dashboard, and its algorithms were based on the Canadian HF guidelines (16,17). Patient’s vital signs (from the mobile app for the RPM^+^/DTx^+^ group or entered manually for the RPM^-^/DTx^+^ one), clinical characteristics (NYHA class, ejection fraction, comorbidities), current medication and most recent blood test results were used as inputs. The output consisted of a tailored report, in which a summary of the patient’s status was displayed and suggestions on how HF therapy could be optimized. This report was generated twice, at day zero and six weeks. It was made available to patients’ healthcare providers, who could use it at their discretion.

### Statistical analysis

SSPSS (version 28, IBM ©) was used for statistical analysis. Results are expressed as means and standard deviations (SD) or as the number of cases and proportions (%), totals and according to groups. Standard deviations were used for normally and non-normally distributed continuous variables. Chi-square tests for test distribution and independent t-tests to compare groups and results were considered statistically significant if P<0.05. Effect size Hedges’ g and Phi or Cramer’s V coefficients were used accordingly due to small sample sizes. (18)

## Results

### Studied population

A total of 52 patients were enrolled in this pilot study and allocated 32 in the RPM^+^/DTx^+^ group, 9 in the RPM^-^/DTx^+^ group and 11 in the RPM^-^/DTx^-^ one (Figure 2). Baseline characteristics are reported in Table 1. Most patients (75%) were in NYHA 2 class, and half had a reduced left ventricular ejection fraction. There were slightly more hypertensive patients and patients with ischemic cardiomyopathy in the RPM^-^ groups than in the RPM^+^ group. Among the RPM^+^ patients, 6/32 patients were helped by a caregiver to manage the application. Compliance with the app was considered high for 15/32 (47%) patients.

**Table 1.**
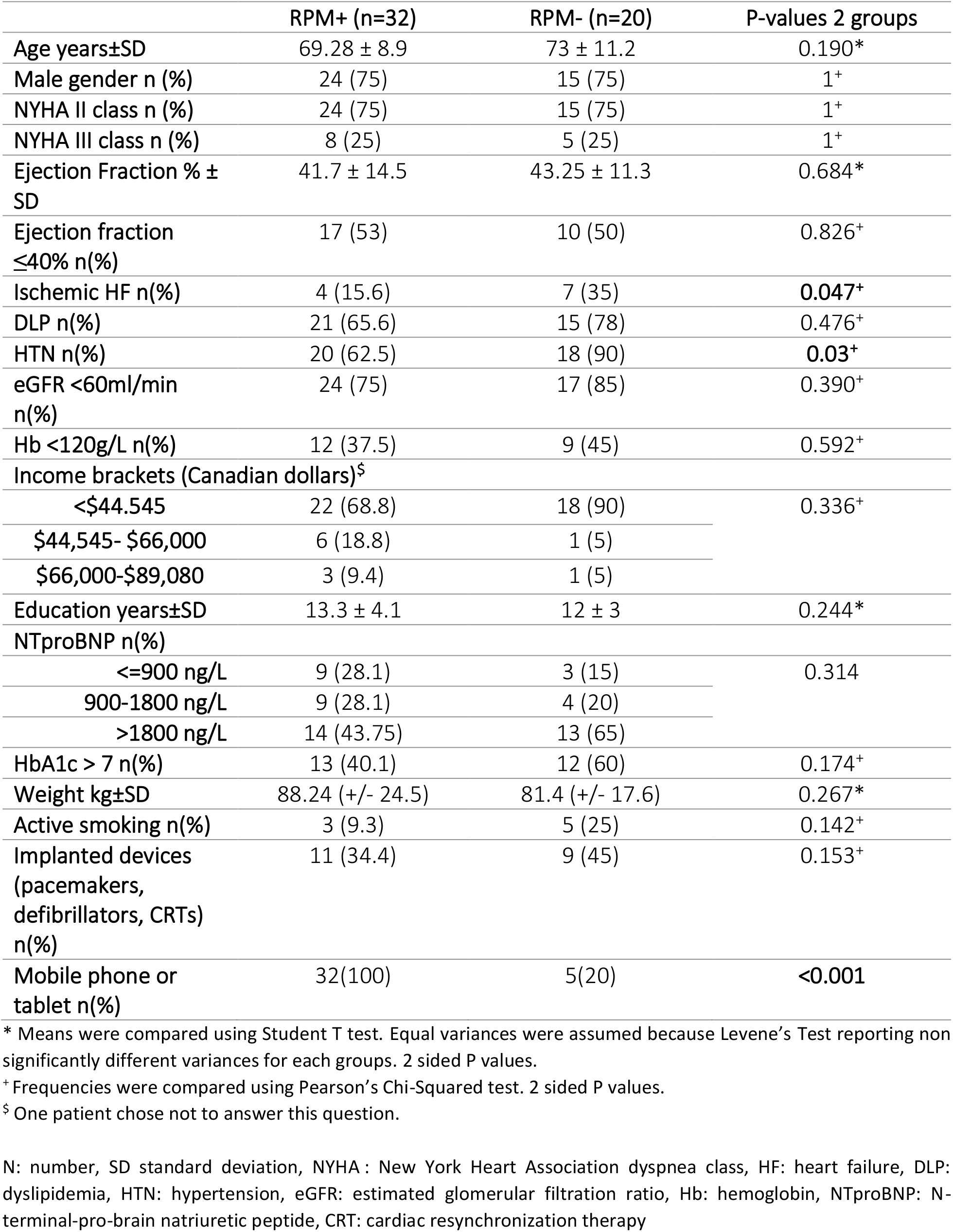
Baseline characteristics

**Figure 2:**
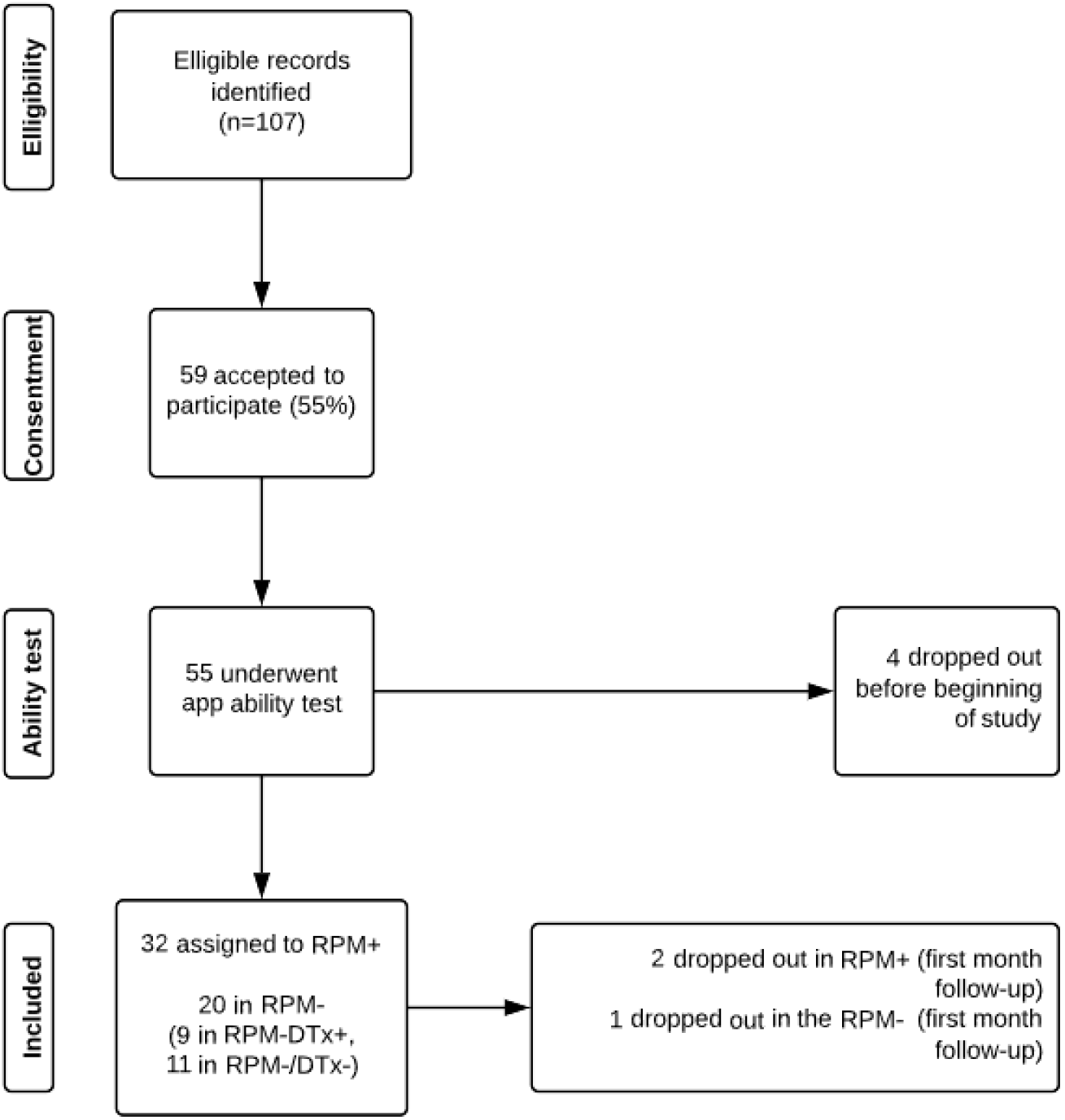
Patient flowchart. RPM+: having remote patient monitoring, RPM-: without remote patient monitoring, RPM-DTx+: without remote patient monitoring and having digital therapeutics, RPM-/DTx-: without remote patient monitoring and without digital therapeutics

### Feasibility

The 15 minutes presentation of the app by the nurse was sufficient for most of the patients, although some required 30 minutes of help. Nurses in charge of the dashboard could integrate those patients’ follow-ups into their regular workday. It usually took them 15-30 minutes per day to review the dashboard for a cohort with at most 30 patients. Alert management, including calls and virtual visits, was performed subsequently if needed. A total of 1322 automated alerts were triggered during the whole study. Most were about blood pressure (38%) and weight (37%); 12% were about blood glucose, 9% about heart failure symptoms, and 4% about heart rate. The same patient could trigger different alerts at the same time if the patient’s condition was unstable (for example, three alerts occurred at the same time for the same patient because of weight gain, worsened dyspnea, and increased edema). Most of these alerts were not considered clinically significant and were rapidly discarded by the nurse. In forty-one situations, the medical file had to be reviewed by the nurse. Thirty-four patients got called by the nurse (meaning approximately the equivalent of 1 call per patient during the 12 weeks of follow-up). Of those, nine had a medication change, two were referred to the emergency room, one was seen as an urgent case at the HF clinic, and one met an electrophysiologist.

### Clinical impact

HF drug optimization in patients with an ejection fraction <40% occurred more frequently in the RPM^+^/DTx^+^ group (figure 3) than in the two other RPM^-^ ones combined (41% vs 20%, medium effect size, *w*=0.26). This effect size was even more significant when comparing only the highly compliant RPM^+^/DTx^+^ patients to all the others (50% vs 20% of optimization rate respectively, medium to large effect size, *w*=0.41).

**Figure 3:**
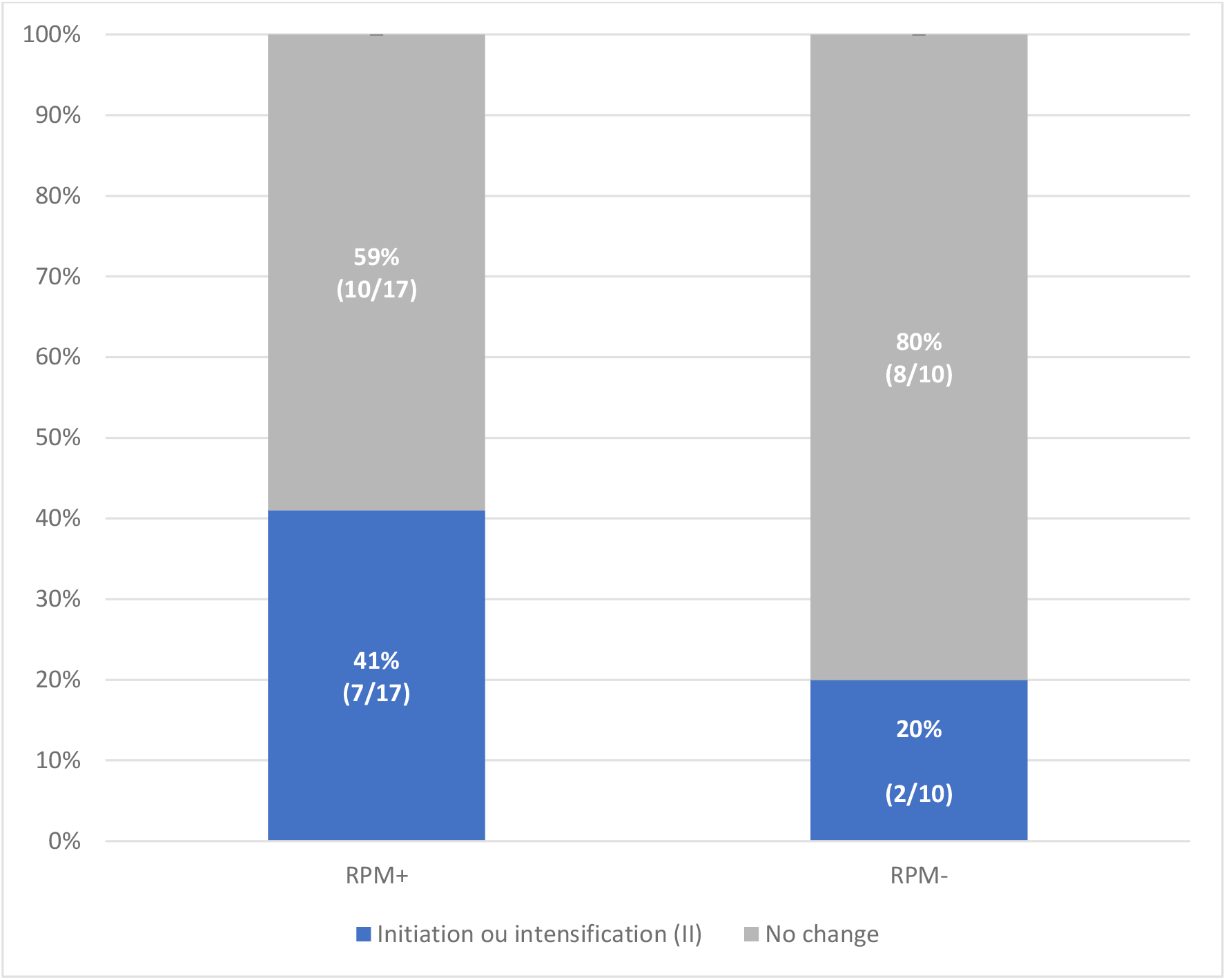
Proportion of patient’s with LVEF ≤ 40 having initiation or intensification of HF medication (at least one class) LVEF: Left ventricular ejection fraction, RPM+: with remote patient monitoring, RPM-: without remote patient monitoring

Quality of life (QOL) assessed by the KCCQ-12 score did not significantly change over the 12 weeks of follow-up, regardless of their group. However, high-compliant RPM^+^/DTx^+^ patients had a trend for a greater KCCQ-12 score at 12 weeks than the other ones (figure 4, medium Hedges’ effect size (g=0.48) after removal of outliers (defined as an increasingly more significant than 1 SD at six weeks and not sustained in the following evaluation)).

**Figure 4:**
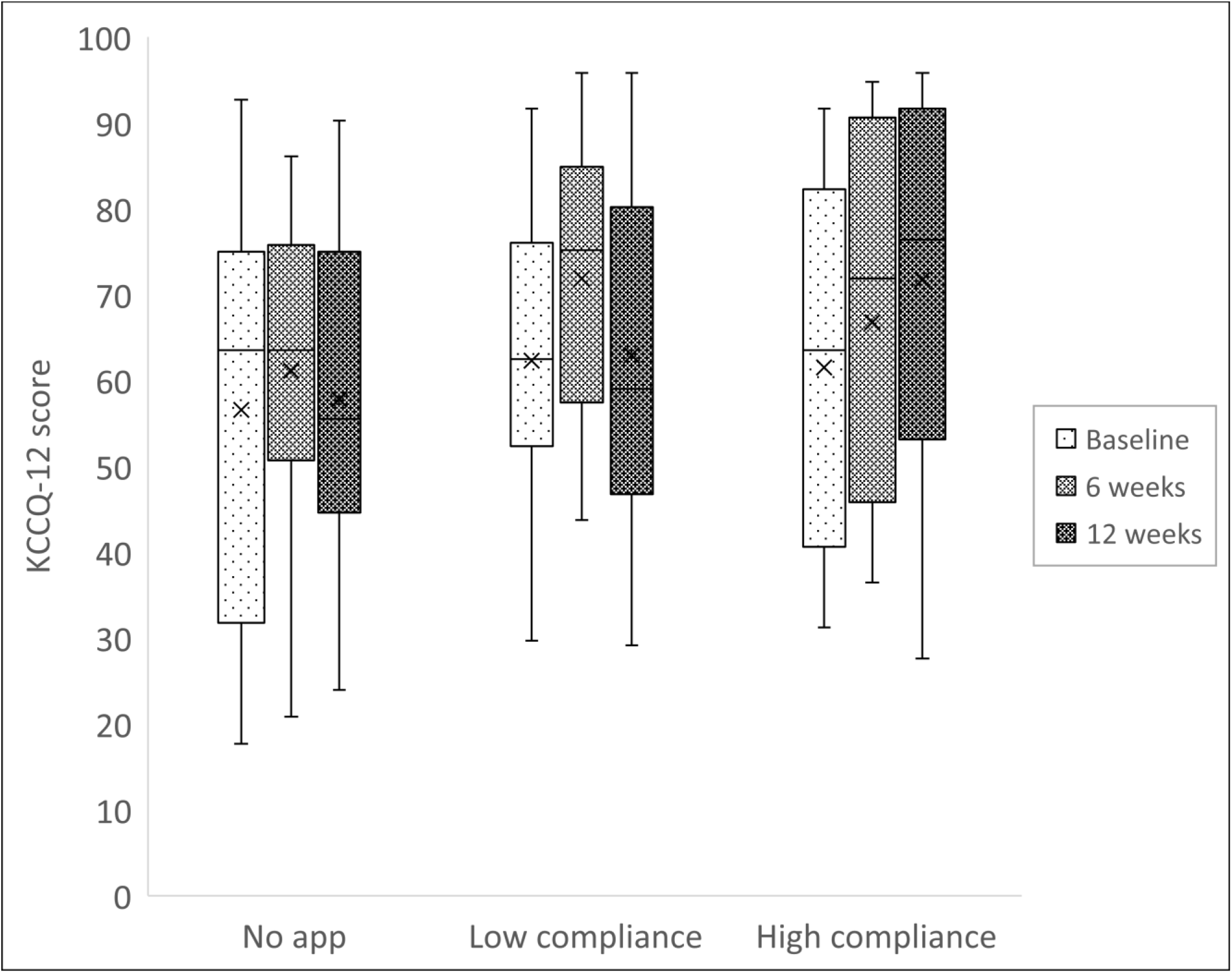
KCCQ-12 Summary Score over time. Hedges’ effect size is medium (g=0.48) after removal of outliers (defined as an increase bigger than 1SD at 6 weeks and not sustained in the following evaluation)).

The number of patients that had at least one hospitalization (all-cause) during the follow-up was much higher in the RPM^-^ groups compared to the RPM^+^ group (35% vs 6%; p=0.008) (Figure 5). Similarly, HF-related hospitalizations tended to be more frequent in the RPM^-^ groups than in the RPM^+^ group (25% vs 6%, p=0.054) (Figure 5).

**Figure 5:**
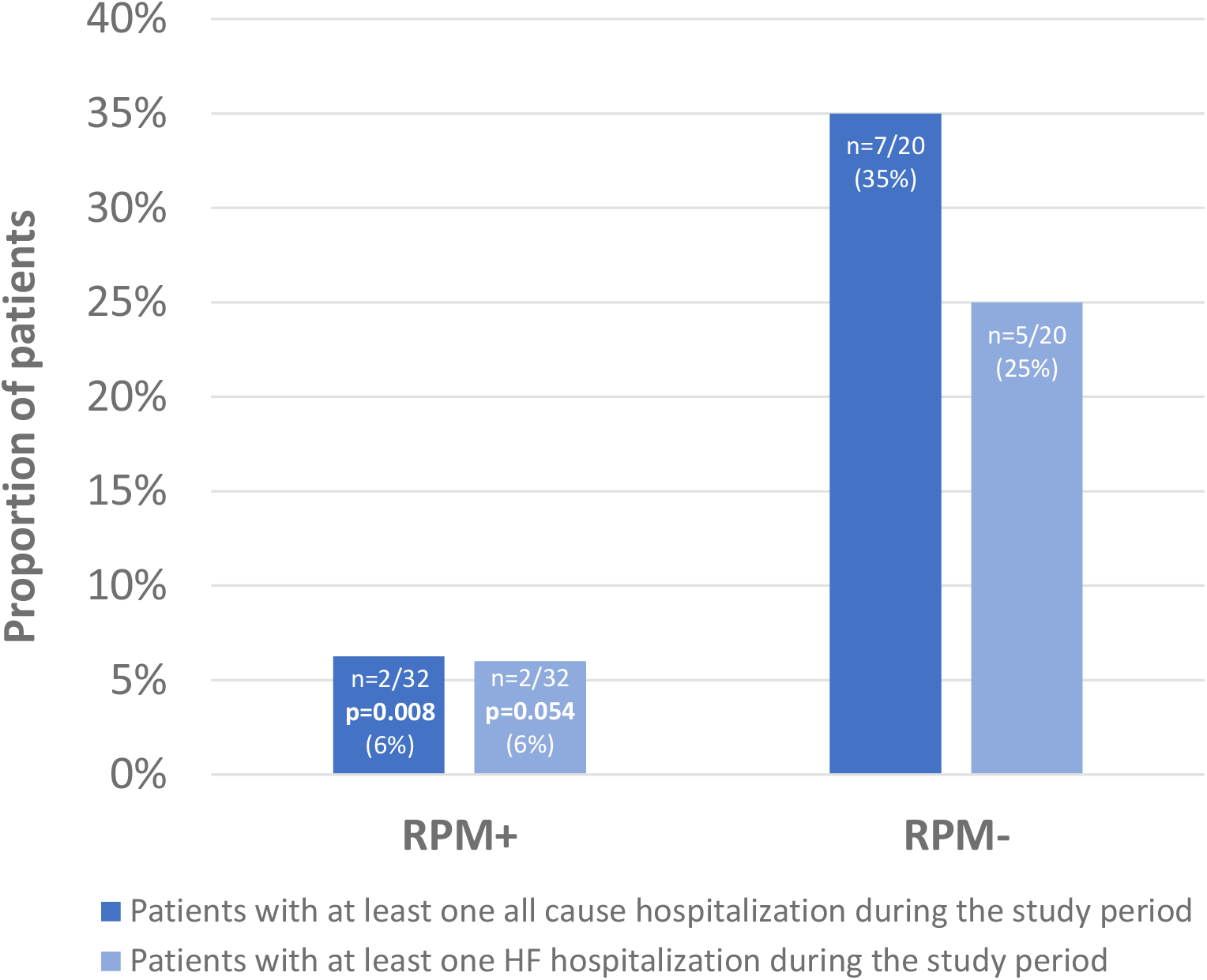
Patients with at least one all cause hospitalization during the study period. RPM+: with remote patient monitoring, RPM-: without remote patient monitoring, HF: heart failure The Chi-square statistic is significant at the .05 level. More than 20% of cells in this subtable have expected cell counts less than 5. Chi-square results may be invalid.

## Discussion

This prospective pilot study demonstrated the feasibility and potential benefits of using a dual solution (RPM + DTx) for HF patients. With this app, they tended to be less hospitalized, have a greater quality of life, and be more likely optimized in terms of HF therapy.

### Technology’s use and implementation

An RPM solution aims to prevent acute HF decompensation and hospitalization, to improve patient engagement and self-care (10,19). Consequently, the Continuum platform and its mobile app have been designed to be user-friendly so healthcare professionals can promote its use and engage patients’ participation. In our study, 47% of the RPM^+^/DTx^+^ patients were considered highly compliant, similar to other studies (compliance ranging from 37-99%). However, compliance definition and technology used might differ from one study to another. (20)

Low income, low level of education, living alone and frailty (21) are barriers to access to these technologies. Nevertheless, mobile phone use has more than doubled in the last decade. (22) Sixty percent of Canadians over 65 have a mobile phone (23). In our study, 62% (32/52) of the patients could rapidly use the app.

RPM implementation’s success is strongly associated with its workflow integration. (24) As available healthcare workers’ resources in Canada are becoming scarce, telemedicine can be one of the solutions to this issue. However, it should not increase healthcare professionals’ workload since telemedicine’s cost-effectiveness has not yet been proven (25) and the growing amount of non-relevant data can constitute a barrier to its implementation. Therefore, thresholds for alerts must be constantly reassessed and adapted (24) to allow professionals to distinguish what is relevant and what is not.

### DTx module: impact on HF drug optimization

Although HF drug optimization is proven to help decrease mortality and morbidity, (2,4,11) this is rarely achieved. (5) DTx can be part of the solution since it can deliver various interventions to treat, manage, or prevent a medical disorder or disease. (14) Our DTx interface focuses on HF therapy but needs efforts to stay updated as new treatments and guidelines evolve. (4,11,16,17)

### Quality of life and hospitalizations

In our study, there was only a trend in favour of Continuum for our highly compliant patients to the intervention in terms of KCCQ-12 scores. It is similar to what others reported. (26) For hospitalization, the RPM-control groups had a rate of 35% all-cause hospitalizations during the three months follow-up. We assumed it might be related to a more vulnerable and frailer subgroup than regular HF patients, as already analyzed.(21)

#### Limitations

Each part of the Continuum platform (the mobile application and RPM and DTx modules) was continuously developed during the study to integrate patient and clinician feedback. It took time to meet clinicians’ expectations. The impact of Continuum on patients enrolled at the beginning of the study versus those enrolled much later could have been significantly different.

## Conclusion

In summary, this pilot study shows that combining RPM and DTx solutions to manage HF patients is feasible and may have some strong positive impact. To our knowledge, it is the first DTx solution developed to help clinicians with HF treatment optimization. This technology is currently being tested in a randomized controlled trial (NCT05377190). (27)

## Data Availability

All data produced in the present study are available upon reasonable request to the authors

## Acknowledgements

We acknowledge the participation of the CHUM hospital workers, including the heart failure clinical nurse’s department (Berraca Jolimeau, Cécile Merceron and Mira Abi-Raad), the contributors in the cardiology department and the telehealth department at CHUM (Rudolph de Patureaux). We also wish to thank our partners at Greybox Solutions Inc. (Pierre Bérubé, Xavier Jodoin and Lydie Montesinos) and Boehringer Ingelheim, Canada Ltd (Suzanne Kimmerle and Eve Blanchet).

## List of declarations

### Research support

This work with the CRCHUM has been supported by a research grant from Medteq^+^ (QC, Canada) in partnership with Prompt (QC, Canada) and Mitacs (QC, Canada) by Boehringer Ingelheim Canada Ltd, and Greybox Solutions Inc. (Montreal QC, Canada).

### Conflict of interest

The first author has received speaker fees from Boehringer Ingelheim Canada, and the last author is the President of the Quebec Heart Failure Society (QHFS).

## Notes

### Clinical Trial

Pilot study #21.403, active RCT following pilot study: NCT05377190

### Author Declarations

Ethics committee of Hospital of the University of Montreal gave ethical approval for this work

